# A two-sex model of human papillomavirus infection: Vaccination strategies and a case study

**DOI:** 10.1101/2021.12.19.21268067

**Authors:** Shasha Gao, Maia Martcheva, Hongyu Miao, Libin Rong

## Abstract

Vaccination is effective in preventing human papillomavirus (HPV) infection. It still remains debatable whether males should be included in a vaccination program and unclear how to allocate the vaccine in genders to achieve the maximum benefits. In this paper, we use a two-sex model to assess HPV vaccination strategies and use the data from Guangxi Province in China as a case study. Both mathematical analysis and numerical simulations show that the basic reproduction number, an important indicator of the transmission potential of the infection, achieves its minimum when the priority of vaccination is given to the gender with a smaller recruit rate. Given a fixed amount of vaccine, splitting the vaccine evenly usually leads to a larger basic reproduction number and a higher prevalence of infection. Vaccination becomes less effective in reducing the infection once the vaccine amount exceeds the smaller recruit rate of the two genders. In the case study, we estimate the basic reproduction number is 1.0333 for HPV 16/18 in people aged 15-55. The minimal bivalent HPV vaccine needed for the disease prevalence to be below 0.05% is 24050 per year, which should be given to females. However, with this vaccination strategy it would require a very long time and a large amount of vaccine to achieve the goal. In contrast with allocating the same vaccine amount every year, we find that a variable vaccination strategy with more vaccine given in the beginning followed by less vaccine in later years can save time and total vaccine amount. The variable vaccination strategy illustrated in this study can help to better distribute the vaccine to reduce the HPV prevalence. Although this work is for HPV infection and the case study is for a province in China, the model, analysis and conclusions may be applicable to other sexually transmitted diseases in other regions or countries.

## 1. Introduction

Human papillomavirus (HPV) is mainly transmitted through sexual contact. There are more than 100 types of HPV, among which at least 14 can cause cancer and are known as high risk types. Almost all sexually active people are infected at some point in their lives and some may be repeatedly infected. HPV infections usually clear up without any intervention within a few months after acquisition, and about 90% will clear within 2 years. However, in some cases, HPV infection can persist and progress to cancer [1]. Cervical cancer is the most common HPV-related cancer in women, with an estimated 569,847 new cases and 311,365 deaths in 2018 globally. There is some evidence linking HPV infection with other cancers of vulva, anus, vagina, penis and oropharynx. In the high risk type group, HPV 16/18 cause 70% of cervical cancers and pre-cancerous cervical lesions. In the low risk type family, HPV 6/11 result in 90% of genital warts and most RRP (recurrent respiratory papillomatosis) [1].

Vaccination is an effective way to prevent HPV infection. There are currently 3 prophylactic vaccines available. The bivalent, quadrivalent and 9-valent vaccine protect people from HPV types 16/18, 16/18/6/11 and 16/18/6/11/31/33/45/52/58, respectively. HPV vaccines have been shown to be safe and very effective in preventing HPV infection and its sequelae [2, 3]. HPV vaccine works more effectively if injected before potential exposure to HPV. Therefore, the World Health Organization (WHO) recommends to vaccinate girls aged 9-14 [1]. By October 2019, more than 100 countries have introduced HPV vaccines to their national schedules [4]. In many countries, HPV vaccines are only offered to pre-adolescent girls (may also include catch-up programs for older females). An increasing number of countries such as Australia, the US and UK also recommend HPV vaccines to pre-adolescent boys and young men, including men who have sex with men (MSM) [5–7]. However, due to the shortage of HPV vaccines, the WHO has called for countries to suspend vaccination of boys in December, 2019 [8]. There has been a long-standing debate about offering HPV vaccines to boys. Without vaccinating boys, health benefits brought from vaccine would not be maximized [9]. However, a few transmission dynamic models showed that strong herd effects were expected from girls-only policy, even with coverage as low as 20% [10]. Should boys be included in a vaccination program? How to allocate HPV vaccines available in a place to maximize the benefit? These questions need to be further investigated.

During the past two decades, a number of mathematical models have been developed to study the epidemiological and economic consequences of HPV vaccination. Elbasha et al. constructed a dynamic model including both demographic and epidemiologic components to assess the epidemiologic consequences and cost-effectiveness of quadrivalent HPV vaccination strategies [11]. They found that vaccinating girls and women was cost-effective and including men and boys was the most effective strategy. Using a similar model, they showed that the quadrivalent HPV vaccines would be cost-effective when administered to females aged 12-24 years or to both females and males before age 12 with a 12-24 years of age catch-up program [12]. By updating the above two models, Elbasha and his collaborator concluded that expanding the current quadrivalent HPV vaccines to boys and men aged 9-26 could provide tremendous public health benefits and was also cost-effective in the United States [13].

The models from ref. [11–13] provided a framework, on which more mathematical model have been developed to study HPV infection and vaccination [14–20]. For example, Insinga et al. found the most effective strategy for quadrivalent HPV vaccines in Mexico was vaccinating 12 years old children plus a temporary 12-24 years old catch-up program covering both sexes [14]. Gender-neutral program was also considered to be a cost-effective choice in France and Italy [17, 21]. On the other hand, there are several studies concluding that girls-only program is more cost-effective [16, 22–25]. For instance, Cody et al. compared different vaccination strategies of 4-valent or 9-valent HPV vaccines in Japan. They found that the most cost-effective strategy was the vaccination program with 9-valent vaccine targeting 12-16 years old girls together with a temporary catch-up program [16]. Similarly, Kim et al. showed that increasing the coverage in girls was more effective and less costly than including boys in a low-resource setting [22]. Damm et al. found that the cost-effectiveness of additional vaccination for boys was highly dependent on the coverage in girls [26].

Some other papers investigated the impact of HPV vaccines and proper vaccine distributions without considering cost-effectiveness [27–45]. Several of them suggested that girls-only policy was better than including boys, at least in the present situation [27–31]. Vaccinating boys was only reasonable if the vaccination coverage for girls was moderate or high [33]. Brisson et al. found that the benefit of vaccinating boys decreased as the coverage in girls increased [30]. However, Azevedo et al. showed that without including men in a vaccination program the disease could only be controlled when more than 90% of women were vaccinated [32]. Muñoz-Quiles et al. constructed a computational network model and found that HPV-related diseases in women would be eliminated within five decades if the vaccine coverage can achieve 75% for both females and males [44]. In addition, Bogaards et al. found that giving vaccines to the gender with the highest prevaccine prevalence would most reduce the prevalence [28]. Waning immunity was also considered to be important, especially when studying persistent HPV infection and its associated cancer incidence [31, 34]. The impact of HPV vaccination was also evaluated in the population of MSM [27, 44–46]. Díez-Domingo et al. showed that MSM would not benefit by the herd immunity effect of vaccinating females [46]. In our previous paper [27], we found that the heterosexual population gets great benefit but MSM only get minor benefit from vaccinating heterosexual females or males. The priority of vaccination should be given to MSM in order to eliminate HPV infection, especially in places that have already achieved high coverage in females.

Most of the above studies obtained the results on vaccine distribution either from cost-effectiveness analysis or numerical simulations, without providing analytical results. In this paper, we will use a two-sex deterministic model to analytically investigate the vaccine distribution strategy. Using the data from Guangxi Province in China as a case study, we will study what strategies would save time and the total amount of vaccines. Specifically, we will mainly address the following questions: 1. Given a fixed vaccine amount, what is the best way to split the vaccines between the two genders to reduce HPV prevalence? 2. In the case study, what is the threshold of vaccine amount needed to eliminate HPV infection? 3. To reduce HPV prevalence to below a certain threshold, how many years and how many total vaccines are needed under different strategies? How to best allocate these vaccines? To answer these questions, we will formulate and analyze the model in Section 2 and 3, respectively. We conduct a variety of simulations on different vaccine distribution strategies in Section 4. Some discussions of the results follow in Section 5.

## 2. Model formulation

In this section, we formulate a two-sex deterministic model to study the transmission of HPV infection in a heterosexually active population. The population is divided into two groups, namely, heterosexual females and heterosexual males (for simplicity, we will use females and males below), and subscripts *f* and *m* are used to denote them. In each group, the population is divided into 3 classes: susceptible (*S* _*k*_), vaccinated (*V*_*k*_) and infected individuals (*I*_*k*_), where *k* ∈ {*f, m* }.

In the absence of vaccination, we assume that humans become sexually active and enter the susceptible compartment *S* _*k*_ with the recruitment rate Λ_*k*_. They leave a compartment at a rate *μ*_*k*_. Susceptible individuals are infected by HPV with the force of infection *λ*_*k*_. Upon infection, the host moves to the infected compartment *I*_*k*_. Infected people can clear infection at a rate *δ*_*k*_. Although natural recovery can provide protection against future infection for many other virus infections, the situation for HPV might be different. Several studies found that HPV reinfection is common for both females and males, even with the same HPV type [47, 48]. Therefore, in this paper we formulate a deterministic model based on the SIS (susceptible-infected-susceptible) structure, which was also used in some other HPV modeling studies such as ref. [25, 37]. We will discuss the potential influence of adopting a different model, e.g. SIR (susceptible-infected-recovered), on our results.

In the model with vaccination, we assume that a fraction (*ϕ*_*k*_) of susceptibles are vaccinated and vaccine-induced immunity does not wane during the sexually active period. Vaccine offers a degree of protection *τ* (0 ≤ *τ* ≤ 1) regardless of gender. Thus, the probability of a vaccinated person getting infected and moving to the infected compartment *I*_*k*_ is 1 − *τ*. We also assume that all infected individuals, vaccinated or not, can clear infection and become susceptible at a rate *δ*_*k*_. The model is described by the following system of ordinary differential equations. A schematic diagram of the model is shown in Figure 1.

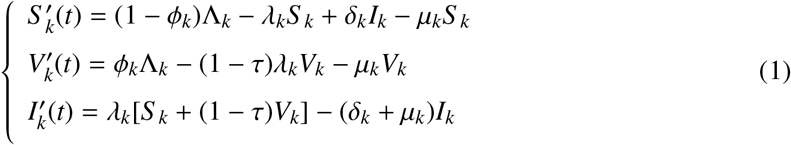

**Figure 1:**
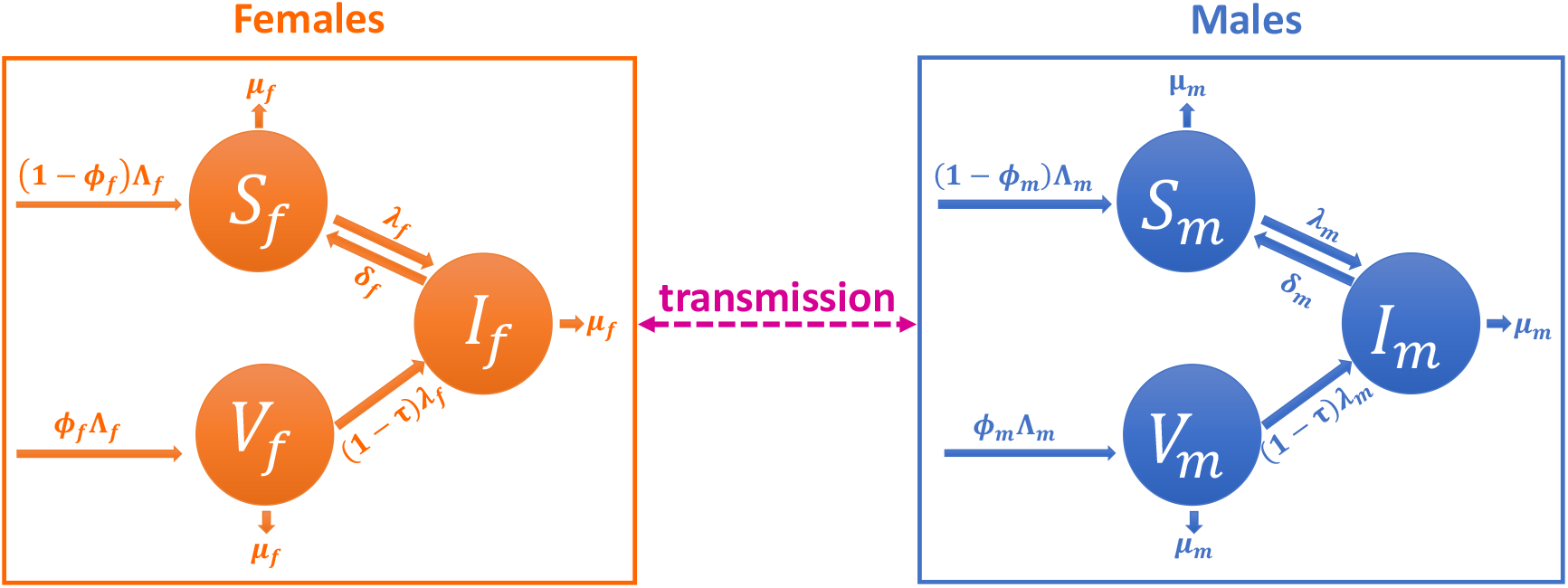
Flow diagram of the model of HPV infection with vaccination. Each group (females and males, denoted by *f* and *m*, respectively) is divided into three subgroups: susceptible, infected and vaccinated, denoted by *S, I* and *V*, respectively. The transmission happens between females and males. Descriptions of parameters are given in Table 1.

The force of infection is given by

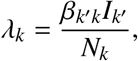

where *N*_*k*_ = *S* _*k*_ + *V*_*k*_ + *I*_*k*_, *k, k ′* ∈ { *f, m*} and *k ≠ k′*. The total population is *N* = *N*_*f*_ + *N*_*m*_.

Taking the sum of *S* _*k*_, *V*_*k*_ and *I*_*k*_ in system (1), we get 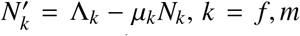. Thus, the equilibrium of *N*_*k*_ is Λ_*k*_/*μ*_*k*_. We define the domain of the system (1) to be

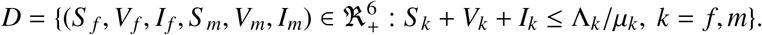

Using a similar method in the previous study [27], we can verify that *D* is positively invariant for system (1) and the model is both epidemiologically and mathematically well posed.

## 3. Analysis of the model

### 3.1. The model without vaccination

The model (1) without vaccination reduces to

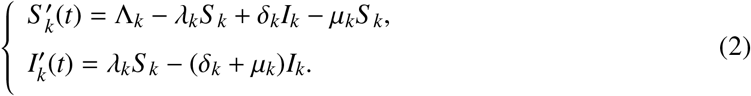

The force of infection is given by

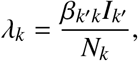

where *N*_*k*_ = *S* _*k*_ + *I*_*k*_, with *k, k′* ∈ {*f, m* } and *k ≠ k*′.

We define

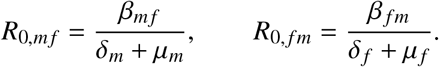

*R*_0,*mf*_ represents the number of secondary female infections generated by one infectious male in an entirely susceptible female population during his whole infectious period. *R*_0, *fm*_ has similar meaning. Using the next generation approach [49], we derive the basic reproduction number to be

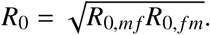

It represents the number of secondary infections generated by one infectious individual in an entirely susceptible population during the whole infectious period of the individual.

The system (2) always has a disease-free equilibrium (DFE) *E*^0^ = (Λ_*f*_ /*μ*_*f*_, 0, Λ_*m*_/*μ*_*m*_, 0). Its local stability is stated in Theorem 1. We present the global stability of the limiting system in Theorem 2. Their proofs are given in Appendix A and B, respectively.

#### Theorem 1.

When *R*_0_ < 1, the DFE *E*^0^ is locally asymptotically stable; when *R*_0_ > 1, the DFE *E*^0^ is unstable.

#### Theorem 2.

When *R*_0_ ≤ 1, the DFE *E*^0^ is globally asymptotically stable.

By setting the right-hand side of system (2) to zero, we can solve for the endemic equilibrium, which is shown in Theorem 3. Its local stability is stated in Theorem 4. The global stability of the limiting system is given in Theorem 5. The proofs of Theorem 4 and 5 are given in Appendix C and D, respectively.

#### Theorem 3.

When *R*_0_ > 1, there exists a unique endemic equilibrium 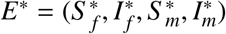, where

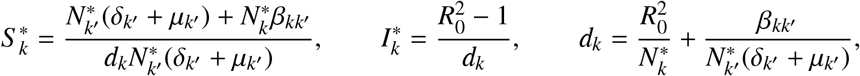

with 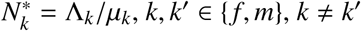.

#### Theorem 4.

When *R*_0_ > 1, the endemic equilibrium *E*^*^ is locally asymptotically stable.

#### Theorem 5.

When *R*_0_ > 1, the endemic equilibrium *E*^*^ is globally asymptotically stable.

### 3.2. The model with vaccination: Analysis and best vaccination strategy

In this section, we study model (1) with vaccination. Similar to the model without vaccination, we define

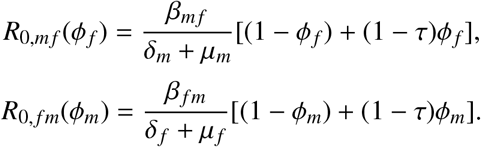

Using the next generation approach, we derive the basic reproduction number to be

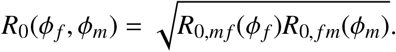

When *ϕ*_*f*_ = *ϕ*_*m*_ = 0, it is the same as the basic reproduction number for the model without vaccination.

The system (1) always has a disease-free equilibrium

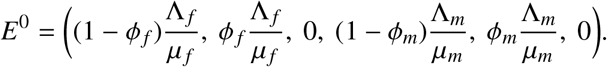

Its local stability is stated in Theorem 6. The proof is similar to Theorem 1 and is omitted. Due to the complexity of the model, it is hard to study the global stability for the DFE. However, using a similar method as in ref. [27], we can show that there is no backward bifurcation for system (1). For the endemic equilibrium, we have the result for its existence, which is stated in Theorem 7 and proved in Appendix E.

#### Theorem 6.

When *R*_0_(*ϕ*_*f*_, *ϕ*_*m*_) < 1, the DFE *E*^0^ is locally asymptotically stable; when *R*_0_(*ϕ*_*f*_, *ϕ*_*m*_) > 1, the DFE *E*^0^ is unstable.

#### Theorem 7.

When *R*_0_(*ϕ*_*f*_, *ϕ*_*m*_) > 1, the endemic equilibrium exists.

From the above analysis, we know that the condition *R*_0_ < 1 is critical for disease elimination. *R*_0_ is also an important indicator that quantifies how fast the disease spreads. Therefore, given a fixed vaccine amount *v*, we investigate the following optimization problem

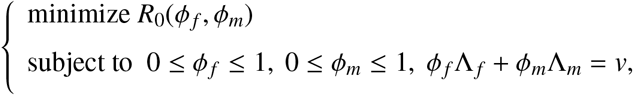

where 0 ≤ *v* ≤ Λ_*f*_ + Λ_*m*_. The result is stated in the following Theorem and the proof is given in Appendix F.

#### Theorem 8.

i. If Λ_*k*_ ≤ Λ_*k′*_, then min *R*_0_(*ϕ*_*f*_, *ϕ*_*m*_) is attained at 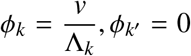 when *v* ≤ Λ_*k*_ and 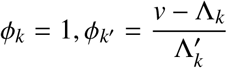 when *v* > Λ_*k*_. Here *k, k ′* ∈ { *f, m*} and *k ≠ k*^*′*^.
ii. When Λ_*f*_ = Λ_*m*_ = Λ, the smaller |*ϕ*_*f*_ − *ϕ*_*m*_|, the bigger *R*_0_(*ϕ*_*f*_, *ϕ*_*m*_). An even distribution (i.e. 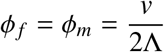) leads to max *R*_0_(*ϕ*_*f*_, *ϕ*_*m*_).

The above Theorem shows that to minimize the basic reproduction number *R*_0_, the gender with a smaller recruit rate should be vaccinated with priority. If there is vaccine left, it will be given to the other gender. When the two genders have the same recruit rate, splitting vaccines evenly is the worst, i.e. resulting in the maximum *R*_0_. The evener the distribution, the bigger *R*_0_.

For convenience, we consider min 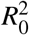 as a function of *v* and denote it by *h*(*v*). We have the following result with the proof given in Appendix G.

#### Corollary 1.

i. *h ′* (*v*) < 0 always holds.
ii. If Λ_*k*_ ≤ Λ_*k′*_, | *h′* (*v*) | is larger when *v* ≤ Λ_*k*_ than that when *v* > Λ_*k*_, where *k, k′* ∈ {*f, m*} and *k ≠ k′*. This Corollary shows that as the vaccine amount *v* increases, the minimum of the basic reproduction number *R*_0_ decreases. Once the vaccine amount exceeds the smaller recruit rate of the two genders, vaccination becomes less effective in reducing the basic reproduction number.

## 4. Vaccination strategies and a case study

### 4.1. Calibration of transmission rates

In this section, we use the data from Guangxi Province in China to calibrate parameter values used in the model. We will investigate various vaccination strategies on the basis of these parameter values. We determine the transmission rates of HPV 16/18 in the model without vaccination. In 2014, an observational cohort study including 2309 men and 2378 women aged 18-55 was conducted in Liuzhou, Guangxi Province (in the end, 1937 men and 2344 women were included in the analysis). Therefore, we set 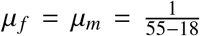. The median time (95% CI) to clear HPV 16/18 is 12.3 [7.7, 13.3] months for females and 6.5 [6.2, 7.7] months for males [50]. So we let the recovery rates (95% CI) for females and males be *δ*_*f*_ = 12/12.3 [12/13.3, 12/7.7] and *δ*_*m*_ = 12/6.5 [12/7.7, 12/6.2] with unit 1/*year*, respectively.

The prevalences of HPV 16/18 for females and males are *p*_*f*_ = 107/2344 = 4.6% and *p*_*m*_ = 30/1937 = 1.5%, respectively [51]. According to Liuzhou Statistical Yearbook [52], there were 2411020 people aged 18-60 in Liuzhou in 2014, and the total female and male populations are 1817723 and 1961637, respectively. Hence the female and male populations aged 18-55 are estimated as

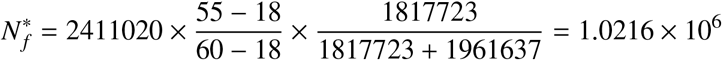

and

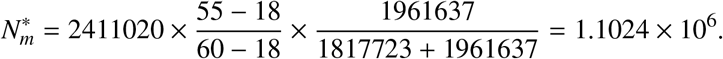

Since HPV infection had existed in Liuzhou for many years before 2014 and HPV vaccine was available until 2016 in mainland China, we assume that HPV infection was in the endemic state at that time. From the expression of the endemic equilibrium of the model without vaccination, we get

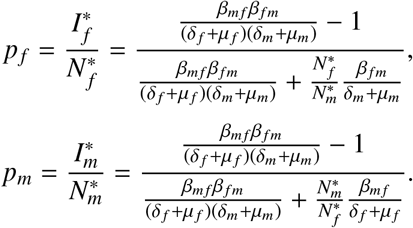

With the above values, we get the transmission rates (95% CI) from females to males is *β*_*mf*_ = 2.8693 [2.6594, 4.5373] and from males to females is *β*_*fm*_ = 0.6967 [0.5896, 0.7299].

### 4.2. Parameter setting

We apply the model to Guangxi province with a wider range of ages 15 − 55. Hence 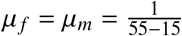. There are two reasons for using a wider age range. One is that some people have sex at young ages (before age 18). The other is that we are interested in the number of target vaccination population (i.e. 14-yea-old children) and want to roughly use it as recruits. The values for *δ*_*f*_, *δ*_*m*_, *β*_*mf*_ and *β*_*fm*_ are the same as above. We estimate the number of 14-year-old boys and girls in Guangxi in 2021-2033 (Figure 2, for details see Appendix H), which are the target vaccination groups. We find that there are only minor changes for both 14-year-old boys and girls from 2021-2033. Therefore, we use the average number as the recruitment rate, namely, Λ_*f*_ = 340998 and Λ_*m*_ = 387463. Applying all these values to the expression of *R*_0_ without vaccination, we derive that the basic reproduction number for HPV 16/18 within the age group 15-55 in Guangxi is 1.0333. Using type-specific and gender-specific clearance rate and prevalence derived from ref. [50, 51], we get basic reproduction numbers for some other HPV types (Table 2).

**Table 1:** Description of variables and parameters.

| Symbol | Description | Baseline | Unit | Range | Source |
| --- | --- | --- | --- | --- | --- |
| **Subscripts** |  |  |  |  |  |
| $f$ | Female | | | | |
| $m$ | Male | | | | |
| $k$ | Gender ($k = f, m$) | | | | |
| **Variables** |  |  |  |  |  |
| $S_k(t)$ | Susceptible population of gender $k$ | | | | |
| $V_k(t)$ | Vaccinated population of gender $k$ | | | | |
| $I_k(t)$ | Infected population of gender $k$ | | | | |
| $N_k(t)$ | Total size of population of gender $k$ | | | | |
| $N(t)$ | Total size of population | | | | |
| $\lambda_k$ | Force of infection for gender $k$ | | | | |
| **Parameters** |  |  |  |  |  |
| $\Lambda_f$ | Recruits into sexually active females | 340998 | person/year | [317422,391244] | See text |
| $\Lambda_m$ | Recruits into sexually active males | 387463 | person/year | [342579,428756] | See text |
| $\mu_f$ ($\mu_m$) | Exit rate from sexually active females (males) | 1/(55-15) | 1/year | See text | |
| $\beta_{mf}$ | Transmission rate from males to females | Calibration | 1/year | Calibration | See text |
| $\beta_{fm}$ | Transmission rate from females to males | Calibration | 1/year | Calibration | See text |
| $\delta_f$ | Recovery rate from infection for females | 12/12.3 | 1/year | [12/13.3,12/7.7] | See text |
| $\delta_m$ | Recovery rate from infection for males | 12/6.5 | 1/year | [12/7.7,12/6.2] | See text |
| $\tau$ | Degree of protection by vaccine | 0.899 | none | [0.817,0.944] | [53] |
| $\phi_k$ | Percentage of new recruits vaccinated for gender $k$ | varied | none | | |
| $v$ | Amount of vaccines per year ($v = \sum_k \phi_k \Lambda_k$) | varied | person/year | | |
| $dummy$ | Parameter for comparison in the PRCC | 1 | none | [1, 10] | See text |

**Table 2:** Calibration for *β*_*mf*_ and *β*_*fm*_ and the corresponding *R*_0_ for different HPV types in people aged 15-55 in Guangxi.

| HPV types | $p_f$ | $p_m$ | $\delta_f$ | $\delta_m$ | $\beta_{mf}$ | $\beta_{fm}$ | $R_0$ |
| --- | --- | --- | --- | --- | --- | --- | --- |
| HPV 16/18 | 107/2344 | 30/1937 | 12/12.3 | 12/6.5 | 2.8693 | 0.6967 | 1.0333 |
| HPV 6/11 | 32/2344 | 23/1937 | 12/6.7 | 12/6.4 | 1.9637 | 1.8068 | 1.0140 |
| HPV 31 | 0.8% | 0.2% | 12/6.5 | 12/6.9 | 6.999 | 0.4775 | 1.0062 |
| HPV 33 | 0.9% | 0.4% | 12/11.7 | 12/6.1 | 2.2147 | 0.9603 | 1.008 |
| HPV 45 | 0.4% | 0.5% | 12/6.7 | 12/6.7 | 1.3532 | 2.4648 | 1.0056 |
| HPV 52 | 6.6% | 1.5% | 12/13.1 | 12/6.9 | 4.1167 | 0.4398 | 1.0443 |
| HPV 58 | 3.5% | 3.5% | 12/12.7 | 12/6.4 | 0.9333 | 2.1271 | 1.0379 |

**Figure 2:**
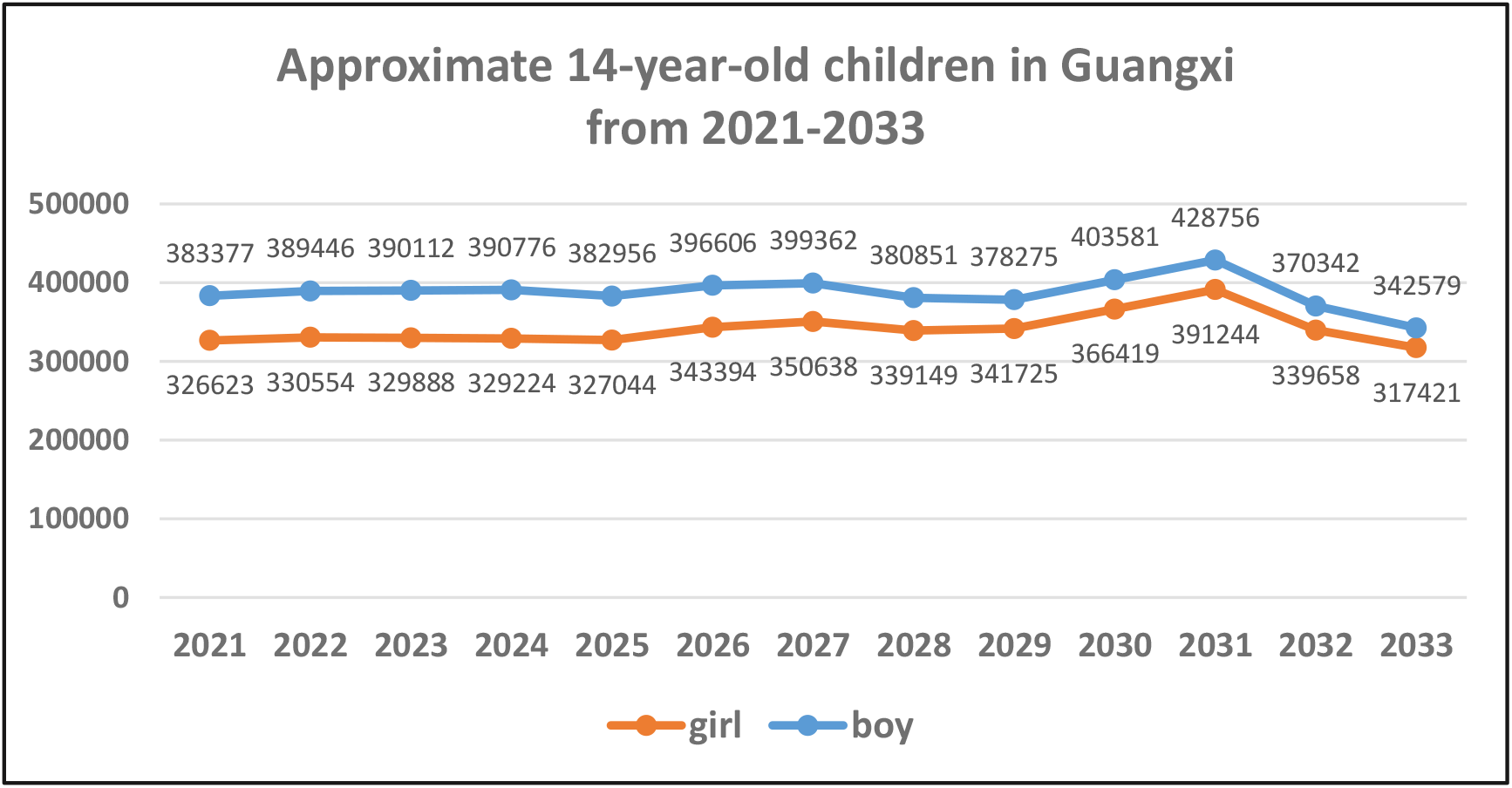
The recruits to susceptible for Guangxi Province. We use data from Liuzhou for parameter calibration and apply to Guangxi. We assume that 14-year-old children are vaccinated and estimate the target population size in 2021-2033 by newborns in 2007-2019.

Chinese domestic bivalent HPV vaccine has been available in Guangxi since 2020. We apply the bivalent HPV vaccine in our model. The value (range) for vaccine efficacy *τ* is 0.899 [0.817-0.944] for HPV 16/18 [53]. So far the bivalent HPV vaccine is only available to females in China. Using all the above values in *R*_0_(*ϕ*_*f*_, *ϕ*_*m*_) and setting *ϕ*_*m*_ = 0 and *R*_0_(*ϕ*_*f*_, *ϕ*_*m*_) = 1, we calculate the critical value of *ϕ*_*f*_ for HPV 16/18 elimination is 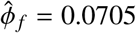. The corresponding vaccine amount is 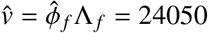.

### 4.3. Sensitivity analysis

We use the partial rank correlation coefficient (PRCC) to evaluate the impact of model parameters on the dynamics of the model (1). The PRCC provides a global sensitivity analysis for nonlinear but monotone relationships between inputs and outputs [54]. From analytical results, we know that *R*_0_ < 1 is essential to eliminate the disease. Therefore, we are concerned with the parameters that have the greatest impact on *R*_0_. We are also interested in the parameters that have great impact on the prevalence. Therefore, in our sensitivity analysis, the inputs are parameters and the outputs are *R*_0_, the prevalence in females, males and the total population. In the inputs, there is a special parameter called dummy, which is introduced to quantify the artifacts (for details see ref. [54]).

In the sensitivity analyses, we choose the vaccination proportion *ϕ*_*f*_ = 0.08 and *ϕ*_*m*_ = 0.01 as baseline values and [0.01, 0.99] as their ranges. Since we only focus on people in the age group 15-55, we fix *μ*_*f*_ = *μ*_*m*_ = 1/(55 − 15) and use one parameter *μ* to represent them. The sample size for *R*_0_ is 10000, and for each prevalence is 6000. The initial condition for the prevalence is pre-vaccination endemic equilibrium, and the end point is 50 years. The negative or positive sign of the PRCC value indicates that the parameter is inversely or positively correlated with the outputs. The parameter with a larger PRCC index (absolute value greater than 0.5) has more significant influence on the output [55]. From Figure 3 and Table 3, we see that the proportions of vaccination *ϕ*_*f*_ and *ϕ*_*m*_ always have a great impact on the basic reproduction number and the disease prevalence. This means that vaccination is an effective way to reduce HPV infection. The output is also sensitive to the clearance rates *δ*_*f*_ and *δ*_*m*_. This indicates that infected individuals having an intact immune response or receiving treatment have a better chance to clear the infection. Other sensitive parameters include transmission rates *β*_*mf*_ and *β*_*fm*_, which highlights the importance of using condom or other protections to reduce the risk of infection.

**Table 3:**
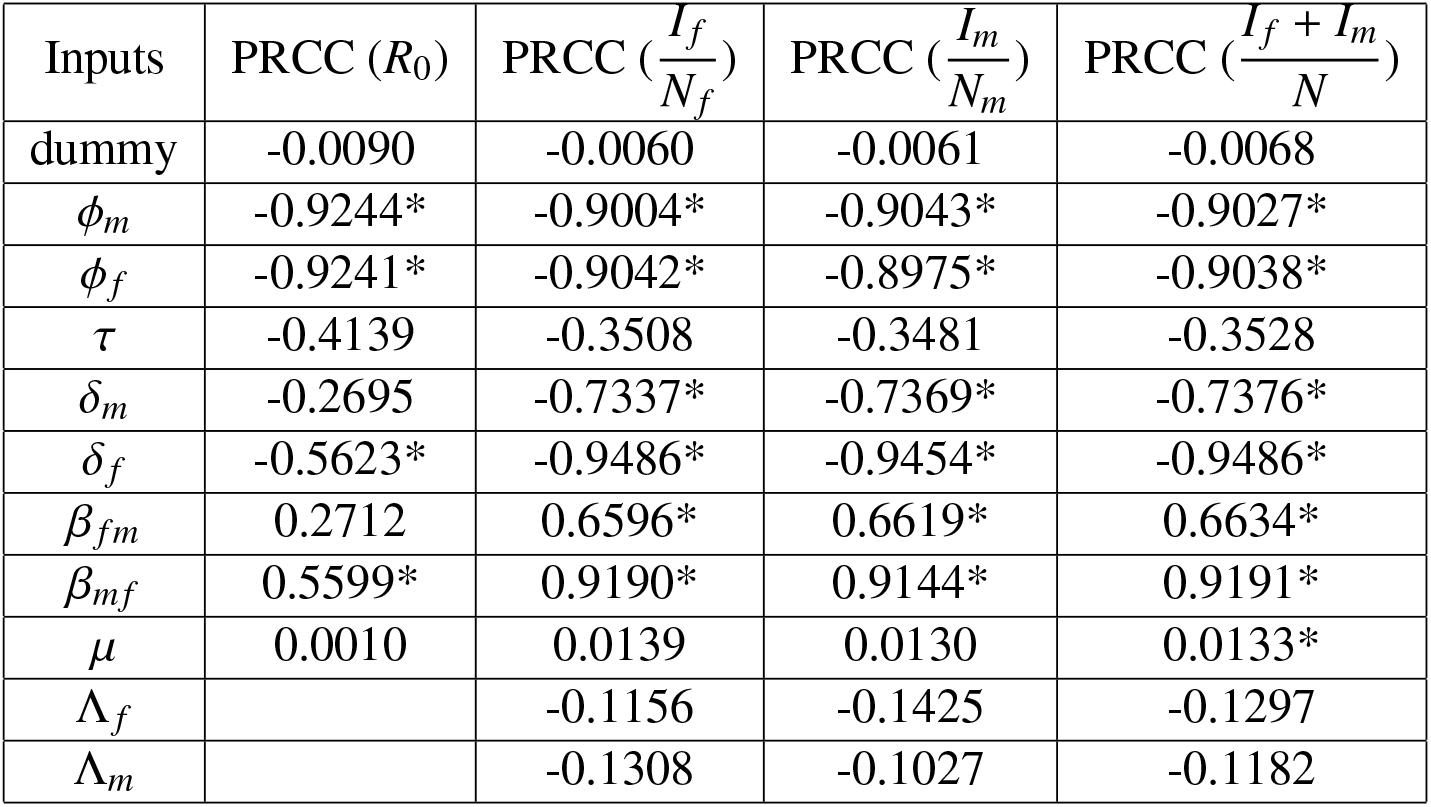
PRCCs for *R*_0_ and prevalences in females, males and the total population (* means significant).

| Inputs | PRCC ($R_0$) | PRCC ($\frac{I_f}{N_f}$) | PRCC ($\frac{I_m}{N_m}$) | PRCC ($\frac{I_f + I_m}{N}$) |
| --- | --- | --- | --- | --- |
| dummy | -0.0090 | -0.0060 | -0.0061 | -0.0068 |
| $\phi_m$ | -0.9244* | -0.9004* | -0.9043* | -0.9027* |
| $\phi_f$ | -0.9241* | -0.9042* | -0.8975* | -0.9038* |
| $\tau$ | -0.4139 | -0.3508 | -0.3481 | -0.3528 |
| $\delta_m$ | -0.2695 | -0.7337* | -0.7369* | -0.7376* |
| $\delta_f$ | -0.5623* | -0.9486* | -0.9454* | -0.9486* |
| $\beta_{fm}$ | 0.2712 | 0.6596* | 0.6619* | 0.6634* |
| $\beta_{mf}$ | 0.5599* | 0.9190* | 0.9144* | 0.9191* |
| $\mu$ | 0.0010 | 0.0139 | 0.0130 | 0.0133* |
| $\Lambda_f$ | | -0.1156 | -0.1425 | -0.1297 |
| $\Lambda_m$ | | -0.1308 | -0.1027 | -0.1182 |

**Figure 3:**
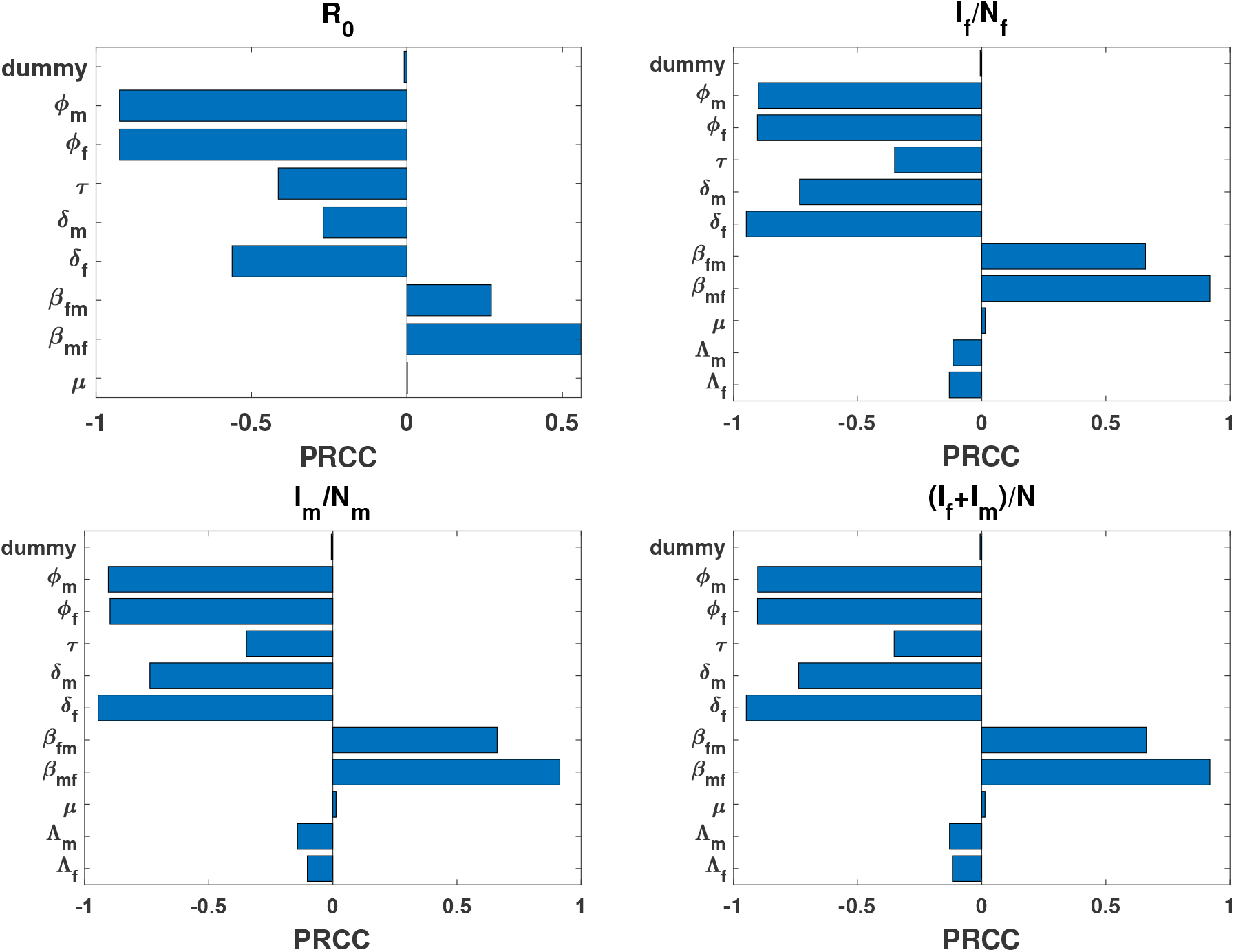
Sensitivity analysis using PRCC. In all panels, we choose 0.08, 0.01 as base values for *ϕ*_*f*_ and *ϕ*_*m*_, respectively, and [0.01, 0.99] as their range. We fix *μ*_*f*_ = *μ*_*m*_ = *μ* = 1/(55 − 15). The other parameters are from Table 1. The initial conditions for (b)-(d) are the pre-vaccination endemic equilibria and the end of time is 50 years.

### 4.4. The best vaccination strategy

The condition *R*_0_ < 1 is critical for disease elimination. We are interested in the minimum vaccine amount needed per year to hit this threshold. Using linear programming, we find that this value is 24050, which is attained when all vaccines are given to girls (Figure 4(a)(b)). It agrees with the results in Section 4.2. To verify that this is a critical value and giving vaccines to girls is better, we set *v* = 25000, which is slightly bigger than the above threshold. We can see min *R*_0_ = 0.9987 < 1, which is obtained when all vaccines are given to girls (Figure 4(c)). In this case all prevalences in females, males and the total population go to zero. They decline faster than when we give all vaccines to boys or split them evenly (Figure 4(d)-(f)). These results are also consistent with the analysis. More specifically, because Λ_*f*_ < Λ_*m*_ in the case study, vaccinating females firstly results in a smaller *R*_0_, which leads to a lower prevalence. Interestingly, even for males, vaccinating girls firstly is still better (see Figure 4(e)). This is reasonable because the transmission always involves both genders. Since Figure 4(d)-(f) show that the dynamics of prevalence in females, males and the total population are similar, from now on we will only focus on the prevalence in the total population.

**Figure 4:**
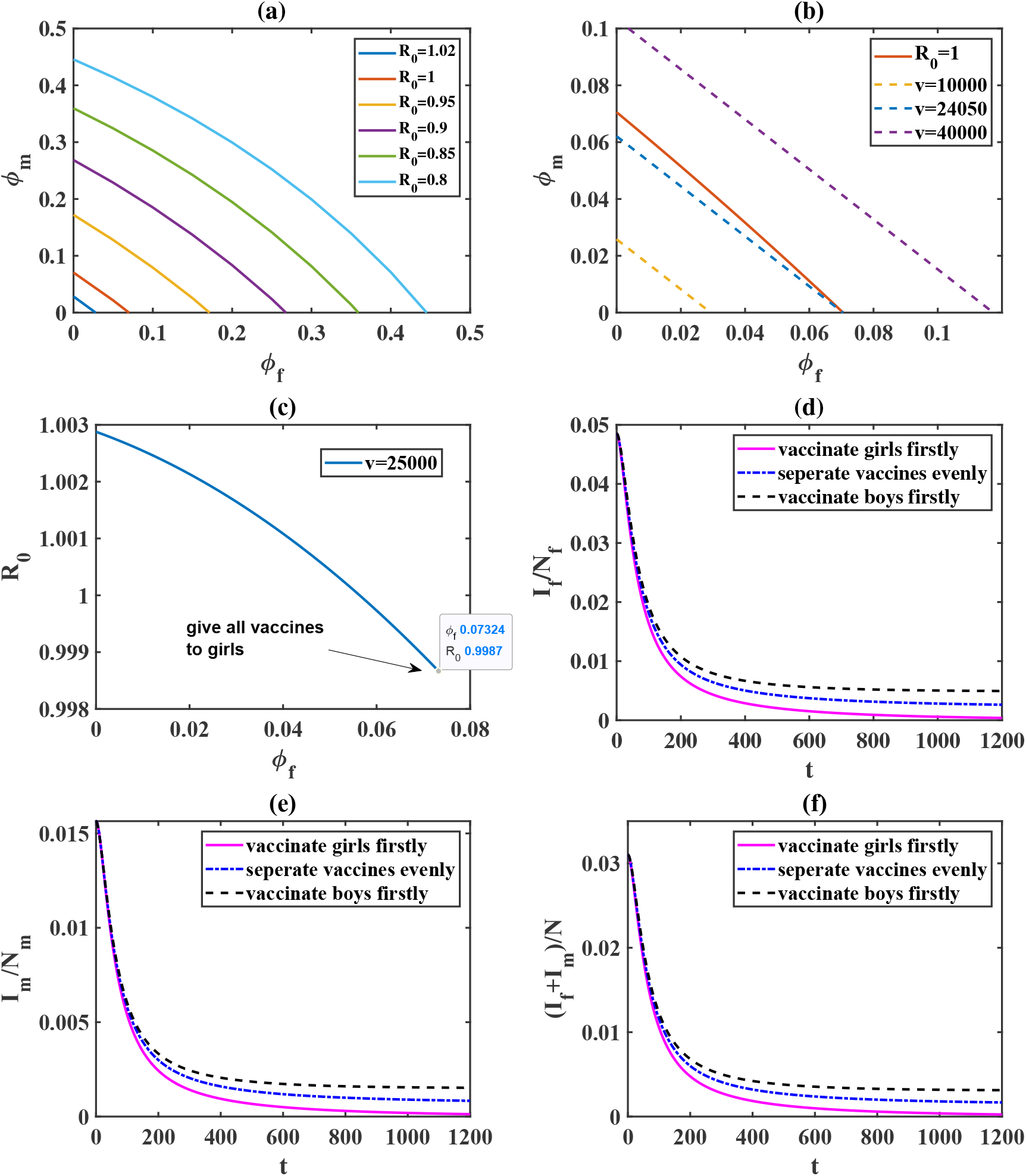
(a) The basic reproduction number *R*_0_ with different vaccine distributions. (b) The minimum vaccine amount v needed per year to achieve *R*_0_ = 1 using linear programming. (c) *R*_0_ for *v* = 25000. The vaccination proportion for males *ϕ*_*m*_ can be calculated according to the vaccine amount *v* and vaccination proportion for females *ϕ*_*f*_. (d-f) Prevalences in females, males and the total population with different vaccine distributions for *v* = 25000. The other parameters are from Table 1. The initial conditions for (d-f) are the pre-vaccination endemic equilibria. It shows minimum *R*_0_ is attained when all vaccines are given to girls, which results in lower prevalences in females, males and the total population.

From Figure 4(f) we also notice that although the disease will go extinct eventually, it will take more than 1000 years and the total vaccine needed will be more than 25000 × 1000 = 2.5 × 10^7^. A critical question arises: can we find a better vaccination strategy requiring less time and less total vaccine amount? Figure 5 shows some vaccination strategies that need less time and less total vaccine amount. Comparing these cases, we estimate that some value *v* ∈ [50000, 150000] will result in the smallest total vaccine amount.

**Figure 5:**
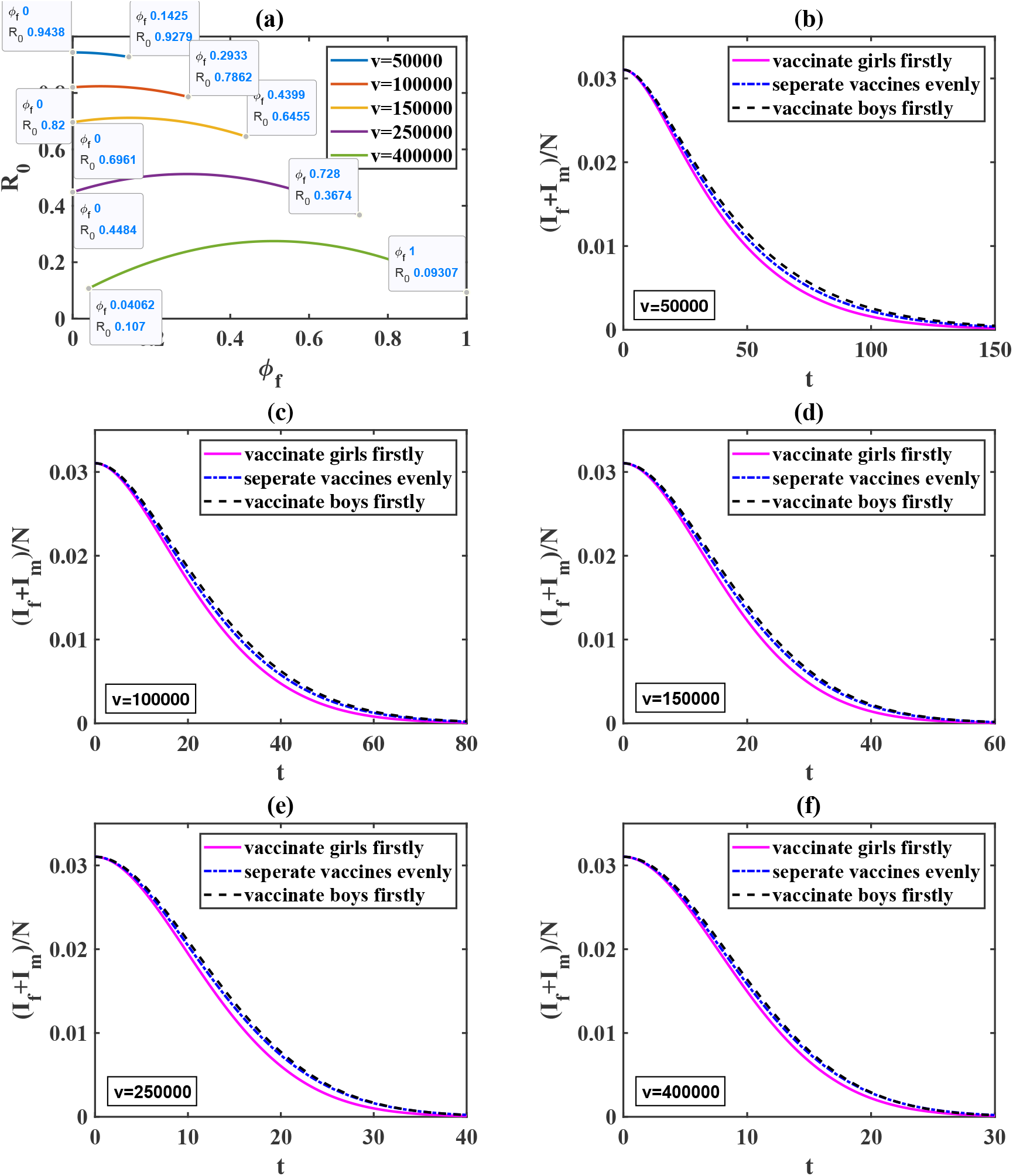
(a) The basic reproduction number *R*_0_ for different vaccine amount *v*. The vaccination proportion for males *ϕ*_*m*_ can be calculated according to *v* and the vaccination proportion for females *ϕ*_*f*_. It shows *minR*_0_ is always attained when girls are vaccinated firstly. (b)-(f) Prevalences in the total population with different vaccine distributions for *v* = 50000, *v* = 100000, *v* = 150000, *v* = 250000, *v* = 400000. The other parameters are from Table 1. The initial conditions for (b)-(f) are the pre-vaccination endemic equilibria.

To further address the above question, we compare the prevalence in the total population for different values of *v* by vaccinating girls firstly (Figure 6). We set a specific threshold 0.0005 for the prevalence in the total population, and compute the total time and total vaccine amount needed to reach this threshold (Table 4). We find that allocating 60000 vaccine shots per year leads to the smallest total vaccine amount (6205014) among these cases. It will take about 103 years to achieve this goal.

**Table 4:** Time and total vaccines needed to reduce the prevalence in the total population to below 0.0005 given fixed vaccine amount every year. All vaccines are given to females. The following vaccine coverage refers to the coverage of 15-year-old girls per year.

| Vaccines (per year) | Vaccine coverage | Time (year) | Total vaccines |
| --- | --- | --- | --- |
| 50000 | $\phi_f=0.15$ | 0-127.3598 | 6367990 |
| 60000 | $\phi_f=0.18$ | 0-103.4169 | 6205014 |
| 70000 | $\phi_f=0.21$ | 0-88.6719 | 6207033 |
| 80000 | $\phi_f=0.23$ | 0-78.2606 | 6260848 |
| 90000 | $\phi_f=0.26$ | 0-70.6311 | 6356799 |
| 100000 | $\phi_f=0.29$ | 0-64.6396 | 6463960 |
| 110000 | $\phi_f=0.32$ | 0-59.8739 | 6586129 |
| 120000 | $\phi_f=0.35$ | 0-55.9049 | 6708588 |
| 130000 | $\phi_f=0.38$ | 0-52.5262 | 6828460 |
| 140000 | $\phi_f=0.41$ | 0-49.8603 | 6980442 |
| 150000 | $\phi_f=0.44$ | 0-47.3095 | 7096425 |

**Figure 6:**
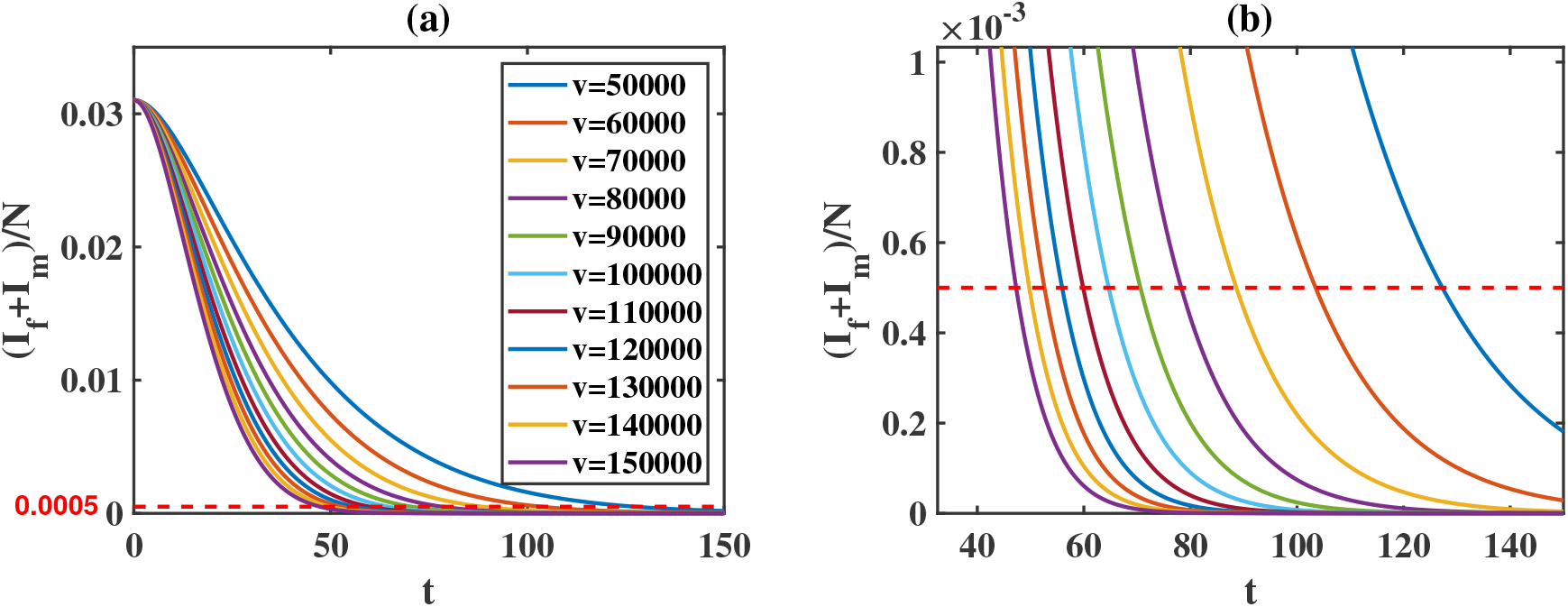
(a) Prevalences in the total population for different vaccine amount *v* assuming vaccinating girls firstly. All vaccines are given to females. The vaccine coverage can be found in Table 4. The other parameters are from Table 1. The initial conditions are the pre-vaccination endemic equilibria. (b) Zoomed figure of the lower part of panel (a).

From Figure 6, we also notice that for any fixed vaccine amount *v*, the prevalence decreases at different speed during different time periods. This indicates that the efficacies of reducing the prevalence with the same amount of vaccine are different. Therefore, we consider variable vaccine amount for different time periods. In Figure 7 (a) we consider three vaccination strategies, namely, adjusting vaccine amount every 50 years, every 30 years and every 20 years. We compare them with the fixed vaccine amount *v* = 60000 per year. Table 5 shows that all the three variable vaccination strategies need less time and less total vaccine amount.

**Table 5:**
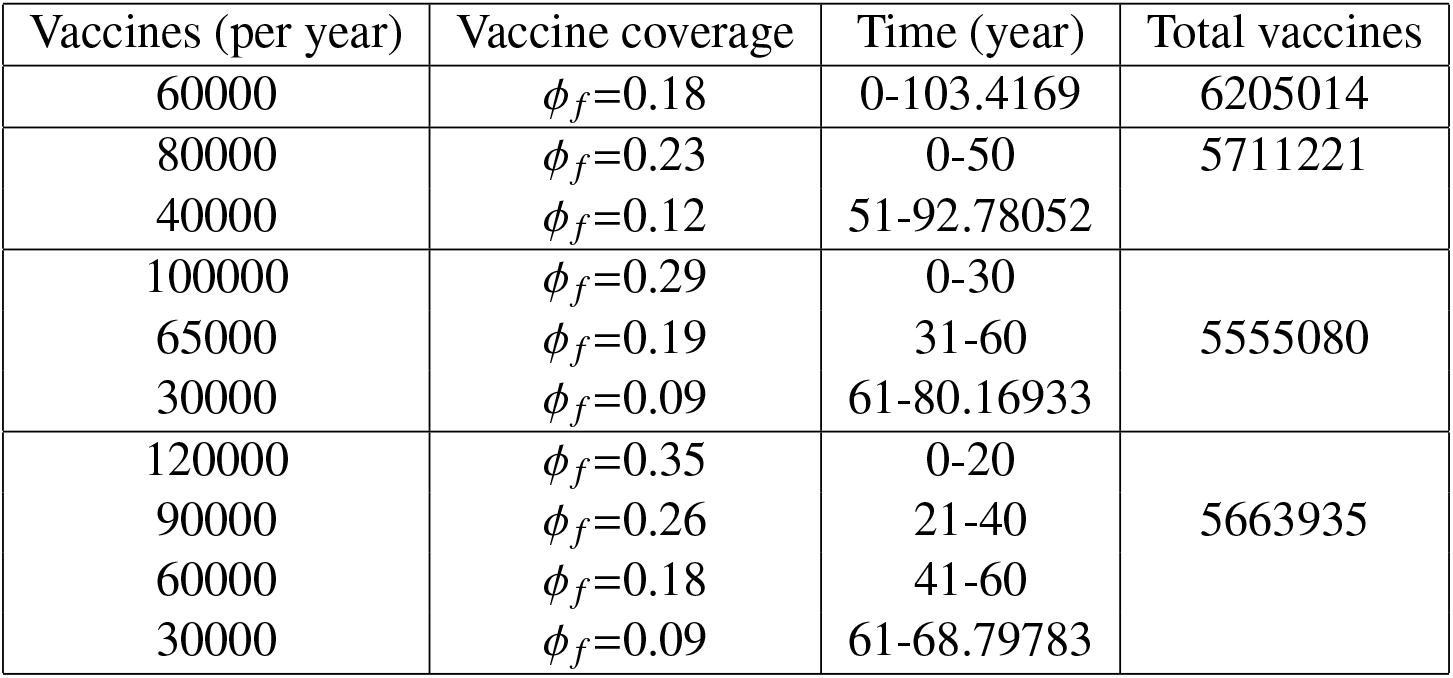
Time and total vaccines needed to reduce the prevalence in the total population to below 0.0005 given different vaccination strategies. All vaccines are given to females. The following vaccine coverage refers to the coverage of 15-year-old girls per year.

**Figure 7:**
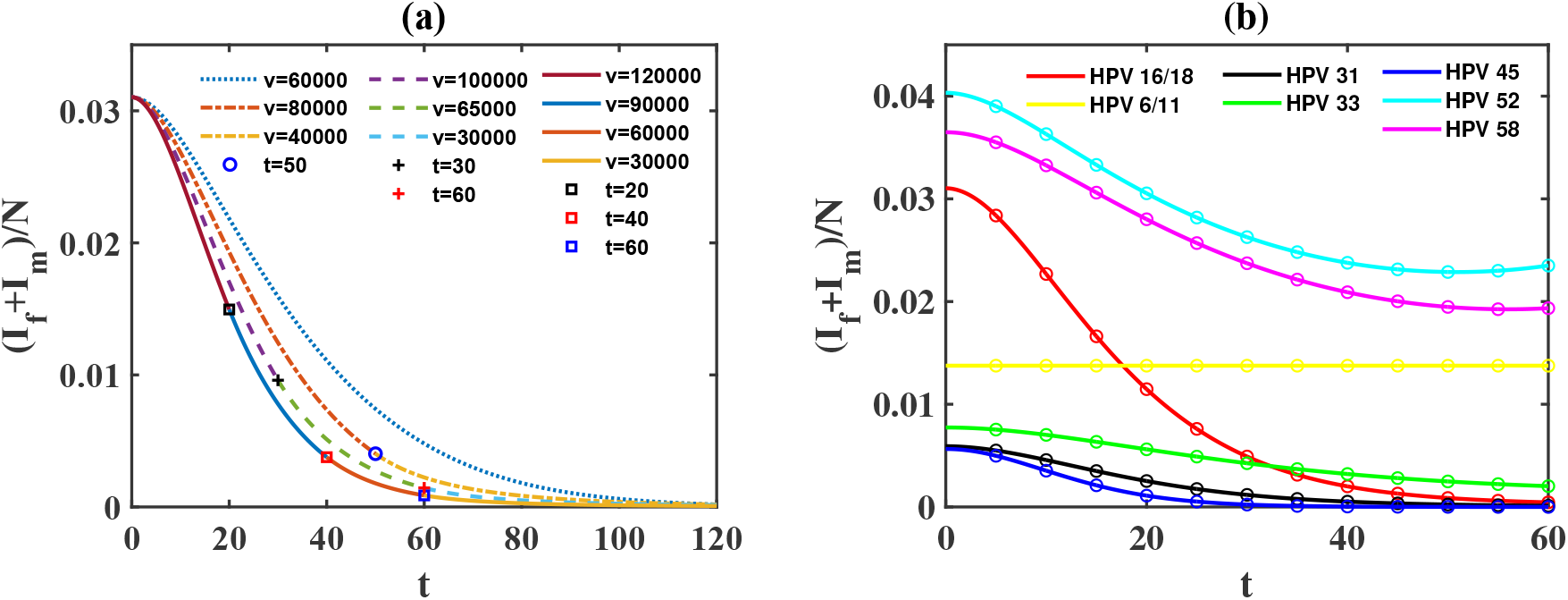
(a) Prevalences in the total population with variable vaccination strategies assuming vaccinating girls firstly. (b) Prevalences in the total population for different HPV types if we allocate vaccine amount *v* = 175000 initially and reduce by 15000 every 5 years assuming vaccinating girls firstly and using bivalent vaccines. According to ref. [53, 66], we let the degree of protection *τ* for HPV 6/11, 31, 33, 45, 52 and 58 be 0, 0.5, 0.257, 0.91, 0.372 and 0.309, respectively. In the two panels, all vaccines are given to females. The vaccine coverage can be found in Table 5 and Table 6, respectively. The other parameters are from Table 1. The initial conditions are the pre-vaccination endemic equilibria.

Furthermore, we consider vaccination with more frequent adjustments. We find strategies with even less time and less total vaccine amount (Table 6). If we allocate 175000 vaccine amount per year in the first 5 years and reduce by 15000 every 5 years, then it only takes about 58 years to achieve our goal and the total vaccine amount is 5225000. If we apply this strategy and consider cross protection of bivalent HPV vaccines for other HPV types, Figure 7 (b) shows the prevalence in the total population for HPV types covered by 9-valent HPV vaccine in the next 60 years.

**Table 6:** Some vaccination strategies with total time less then 68 years and total vaccines less than 5.5 million in order to reduce the prevalence in the total population to below 0.0005. All vaccines are given to females. The following vaccine coverage refers to the coverage of 15-year-old girls per year. The integer *n* = 1, 2, · · · represents the number of period.

| Period length<br>(year) | Vaccines in the first<br>period (per year) | Vaccine<br>deduction | vaccine coverage | Total time<br>(year) | Total<br>vaccines |
| --- | --- | --- | --- | --- | --- |
| 10 | 150000 | 25000 | $\phi_f=0.44-0.07n$ | 67.125 | 5250000 |
| 5 | 175000 | 15000 | $\phi_f=0.51-0.04n$ | 58.0625 | 5225000 |
| 2 | 178000 | 6000 | $\phi_f=0.52-0.02n$ | 59.225 | 5382950 |

Now we consider another case in which there are sufficient HPV vaccines. How to reduce the prevalence in the total population as soon as possible? If we only consider vaccinating adolescent girls, which is the main target population recommended by the WHO, the best situation is that all girls are vaccinated before age 15, namely, *ϕ*_*f*_ = 1 and *ϕ*_*m*_ = 0. In this case, it will take about 27 years to reduce the prevalence in the total population to below 0.0005 (Figure 8 (a)) and the total vaccine amount is 9231805. For a fixed vaccine amount, although vaccinating girls firstly is better, vaccinating boys is still helpful in reducing the prevalence. For example, if we vaccinate all 14-year-old girls and boys every year, then it will take about 19 years to reduce the prevalence in the total population to below 0.0005 (Figure 8 (a)). However, the total vaccine amount is much higher (13895612) in this case. In practice, it is not realistic to achieve 100% coverage. Therefore, we consider more cases with different coverages in Table 7 and Figure 8 (a). We see that a higher vaccine coverage in females and males leads to a smaller *R*_0_ and it takes a shorter time to reduce the prevalence to below 0.0005.

**Table 7:** The values of *R*_0_ with different vaccine distributions.

| $\phi_f$ | $\phi_m$ | $R_0$ |
| --- | --- | --- |
| 0 | 0 | 1.0333 |
| 0.25 | 0 | 0.9098 |
|  | 0.125 | 0.8572 |
|  | 0.25 | 0.8011 |
| 0.5 | 0 | 0.7667 |
|  | 0.25 | 0.675 |
|  | 0.5 | 0.5688 |
| 0.75 | 0 | 0.5897 |
|  | 0.375 | 0.4802 |
|  | 0.75 | 0.3366 |
| 1 | 0 | 0.3284 |
|  | 0.5 | 0.2436 |
|  | 1 | 0.1044 |

**Figure 8:**
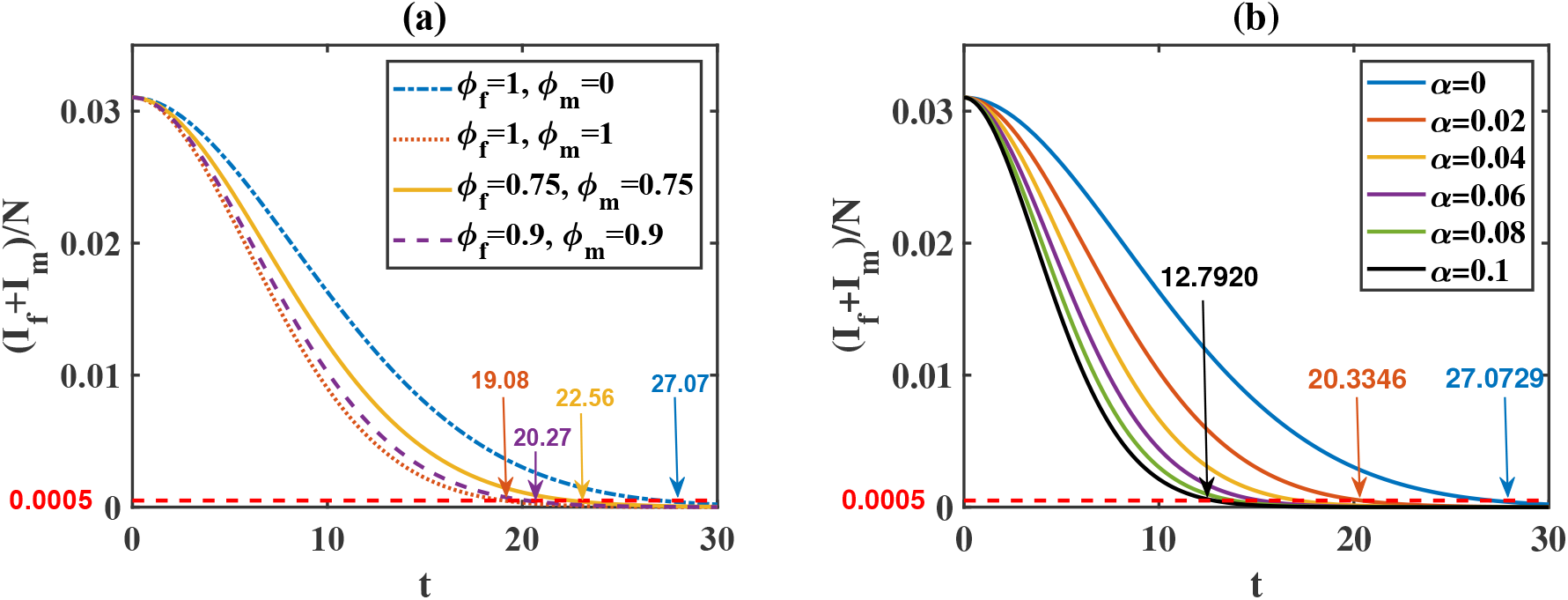
(a) Prevalences in the total population for different vaccination strategies. (b) Prevalences in the total population for different catch-up vaccination proportions with all girls vaccinated by age 15 (*ϕ*_*f*_ = 1, *ϕ*_*m*_ = 0). The other parameters are from Table 1 and the initial conditions are the pre-vaccination endemic equilibria.

Catch-up vaccination (i.e. vaccinate older people) is also useful in reducing the prevalence, especially in the beginning when HPV vaccines are introduced. For example, we assume that *α* is the proportion of women getting catch-up vaccination. Combining with vaccinating all adolescent girls before they become susceptible, the model predicts that the time needed to reduce the prevalence in the total population to below 0.0005 is 20 years when *α* = 0.02 and 13 years when *α* = 0.1 (Figure 8 (b)).

## 5. Discussion

In this paper, we developed a two-sex deterministic model to evaluate the epidemiological impact of HPV vaccination in a heterosexually active population. We derived the basic production number *R*_0_ and studied the stability of equilibria. The analysis shows that *R*_0_ plays a critical role in predicting how the infection spreads. The smaller *R*_0_, the lower prevalence in the total population. In order to reduce *R*_0_, given a fixed vaccine amount, we investigated how to allocate vaccines between the two genders. By rigorous mathematical analysis, we found that min *R*_0_ is achieved when vaccinating the gender with a smaller recruit rate firstly and giving the remaining vaccines, if left, to the other gender. This was numerically illustrated by a case study in which there are fewer female recruits in Guangxi. Vaccinating girls firstly results in the smallest *R*_0_ and lowest prevalence in the total population compared to other strategies. We also considered a special case in which the recruits for females and males are the same. In this case, besides the same conclusion for min *R*_0_, we also found allocating vaccines evenly leads to max *R*_0_. The evener the distribution, the bigger *R*_0_, consequently the higher prevalence. We also proved that min *R*_0_ decreases as the vaccine amount *v* increases but vaccination becomes less effective once its amount exceeds the smaller recruit rate.

The above conclusion on vaccine distribution is reasonable because given a fixed vaccine amount, vaccinating the gender that has a smaller recruit rate enables a larger proportion of the population of that gender getting vaccinated. This leads to more reduction in *R*_0_ and the prevalence in the total population. In a previous paper studying vaccination in a heterosexual population and MSM [27], we assumed the same recruit rate for heterosexual females and males. We found that vaccinating either gender firstly leads to the same prevalence in the total population and this prevalence is lower than splitting vaccines evenly. This agrees with the conclusion here. Bogaards et al. found that vaccinating the gender with a higher pre-vaccination prevalence would result in a larger reduction of the population prevalence [28]. This is also consistent with our result as a higher pre-vaccination prevalence means more people had already gained immunity. Thus, keeping vaccinating that gender will result in a larger proportion of people getting protected. Some other papers also suggested that increasing the vaccine coverage in girls was better than including boys [29–31]. In other words, they also aimed to get a bigger proportion of people protected within the same gender. This strategy indicates that high coverage in one gender can provide strong herd immunity for sexually transmitted diseases, which also agrees with the results in refs. [44, 45].

Using data from Liuzhou, a city in Guangxi province, we calibrated the transmission rates *β*_*mf*_ and *β*_*mf*_. We calculated the basic reproduction number *R*_0_ = 1.0333 for HPV 16/18 in people aged 15-55. Using the estimated recruits in Guangxi, we predicted that the minimum number of bivalent HPV vaccine shots is 24050, which should be given to girls. We also derived basic reproduction numbers for some other HPV types. There are few studies in the literature providing estimates of the basic reproduction number for other HPV types. Riesen et al. estimated that *R*_0_ for HPV 16 in Switzerland was 1.29 [56], and Ribassin-Majed et al. calculated *R*_0_ for HPV 6/11 in France to be 1.04 [37]. The difference in these estimates could be caused by different assumptions. For example, we adopted the gender-specific clearance rate from data, and the other two models used the same clearance rate for both genders. Social and sexual behaviors may also affect the disease spread. Ziyadi et al. obtained a very small basic reproduction number (*R*_0_ = 0.2346) for the African American population [38]. One possible explanation is that the authors used the fitted recruitment and relatively small infection rates.

Although HPV infection is predicted to go extinct once the vaccine amount exceeds a critical value, it would take a very long time and the total vaccine amount could be huge. To find a better vaccination strategy, namely, within a shorter period of time and with less total vaccine amount, we set a specific goal, i.e., the prevalence in the total population is less than 0.05%. We compared several strategies and found that the one offering 60000 vaccine shots to girls every year resulted in the smallest total vaccine amount. In this case, the total vaccine amount was 6205014 and it would take about 103 years to achieve the goal. In addition, we found that the efficacy for the same amount of vaccine was different during different periods. Therefore, the variable vaccination strategies were further studied. We compared several cases, among which the best one was giving 175000 vaccine shots to girls per year in the first 5 years, followed by reducing by 15000 every 5 years. It only took about 58 years to achieve the goal and the total vaccine amount was 5225000. Based on these simulations, it is better to allocate more vaccines at the beginning, and then gradually reduce vaccine shots. Similar results were found in another work [25]. They suggested that vaccination should be applied at the maximum level and after approximately half of the time interval, the rate of vaccination should be gradually reduced, reaching zero in the end. In comparison with the continuous optimal control in [25], our work offered some discrete-time control strategies, which would be easier to implement.

In this paper, we employed a simple two-sex deterministic model to evaluate the epidemiological influence of HPV vaccination and used Guangxi as a case study. The model can be extended to take into account more factors, such as age, sexual behavior and some other heterogeneous mixing. In particular, models with age structure would be more realistic to study the spread of sexually transmitted diseases such as HPV infection and its associated diseases due to the change of sexual behavior with age [44, 45]. We have formulated two such models with age structure to study the dynamics of HPV-associated oropharyngeal cancer and cervical cancer in Texas [19, 20]. The models divide the population into 23 age groups, which is challenging, if not impossible, to obtain any formal analytical results. More comprehensive models would also require more data for parameterization. Here we used the model with a minimum number of parameters that can be calibrated by the case study, performed both analytical and numerical investigations, and provided some quantitative information on the vaccine distribution strategies.

Vaccination of the MSM population might be crucial for disease elimination. Using a model based on a network paradigm and data from Spain, Díez-Domingo et al. found that the MSM group only benefits from a vaccination program that includes males [46]. From both the analysis and numerical investigations in the ref. [27], we showed that in order to eliminate HPV infection, the priority of vaccination should be given to MSM. Because MSM only account for a small portion of the total population, in this paper we focused on the heterosexual population and studied how to allocate HPV vaccine among them. Analysis of the model without MSM provides some analytical results on the vaccine distribution (Theorem 8 and Corollary 1), which are difficult for the full model with MSM. When the MSM population is included, numerical results would suggest similar predictions as in the previous paper [27]. For example, the priority of vaccination should be given to MSM for disease elimination. The heterosexual population gets great benefit but MSM only get minor benefit from vaccinating heterosexual females or males. The best vaccination strategy is to vaccinate MSM firstly as many as possible, then distribute the remaining to the heterosexual population.

We used the SIS model to study the vaccination strategies in view of HPV reinfection after recovery in both males and females [47, 48]. If we use the SIR or SIRS model, the basic reproduction number *R*_0_ remains the same and the prevalences of the male, female and total population show similar dynamics. Therefore, our conclusions are not affected by the choice of these models. In addition, we used the HPV prevalence as a criterion in evaluating various vaccination strategies. If the objective is to reduce HPV-associated diseases such as cervical or oropharyngeal cancer, then the progression from persistent HPV infection to these cancers should be incorporated into models [19, 20, 31, 57–59] and the guideline of vaccination might be different from that informed by this study. Lastly, the parameters and predictions are based on a case study in Guangxi Province in China. This can be applied to other countries or regions. The vaccination strategies obtained in this study may also be applicable to other sexually transmitted diseases.

## Data Availability

All data produced in the present work are contained in the manuscript

## Appendix A Proof of Theorem 1

Reordering variables as *x* = (*I*_*f*_, *I*_*m*_, *S*_*f*_, *S* _*m*_)^*T*^, the Jacobian matrix of system (2) evaluated at the disease-free equilibrium 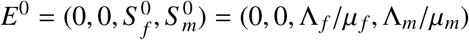 is

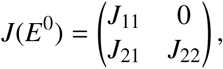

where

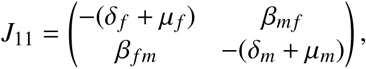

and

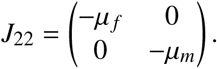

Clearly, −*μ*_*f*_ and −*μ*_*m*_ are negative eigenvalues of *J*(*E*^0^). The remaining eigenvalues are determined by the matrix *J*_11_. If *R*_0_ < 1, then we have *Tr*(*J*_11_) < 0 and 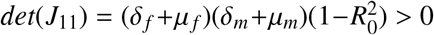. By the Routh-Hurwitz criterion [60], the DFE *E*^0^ is locally asymptotically stable.

If *R*_0_ > 1, then *det*(*J*_11_) < 0. Hence *J*_11_ has an eigenvalue with positive real part. This shows that *E*^0^ is unstable.

## Appendix B Proof of Theorem 2

Considering the limiting system of (2), i.e. when 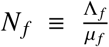 and 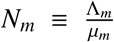, we define the following Lyapunov function

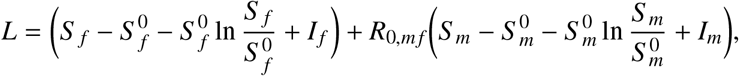

where 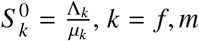. It is clear that *L* is radially unbounded and positive definite in the entire space *D*. The derivative of *L* along the trajectories of system (2) yields

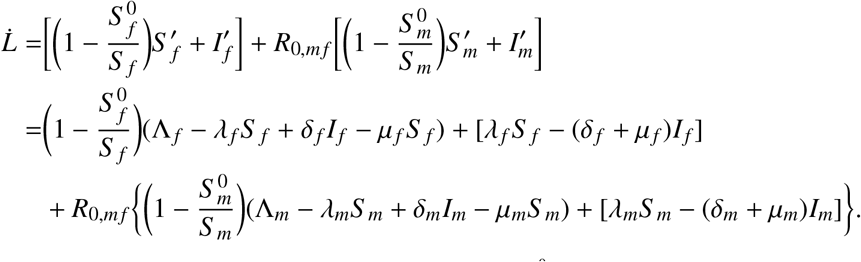

Using the equilibrium conditions 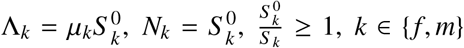 and collecting terms, we obtain

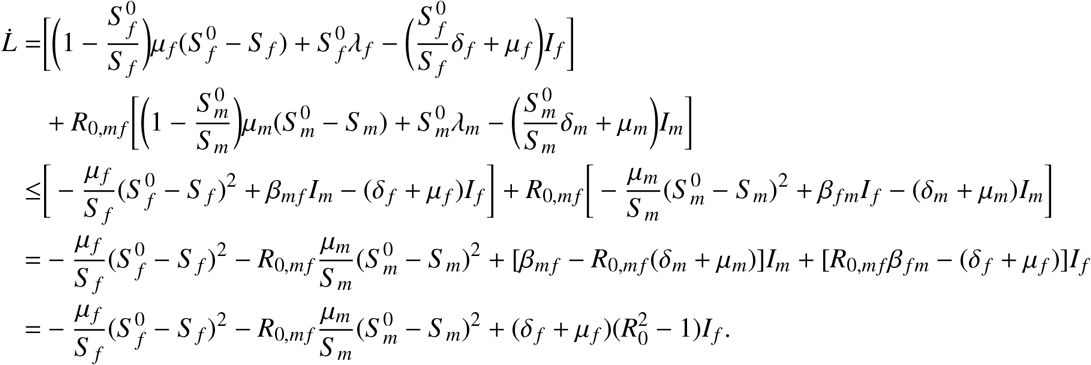

When *R*_0_ < 1, we have that 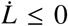 and it is equal to 0 only at the DFE. Therefore, by Krasovkii-LaSalle Theorem [61], the DFE *E*^0^ is globally asymptotically stable when *R*_0_ ≤ 1.

## Appendix C Proof of Theorem 4

The Jacobian matrix of system (2) evaluated at the endemic equilibrium 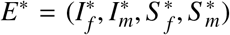 is

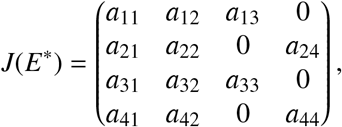

where

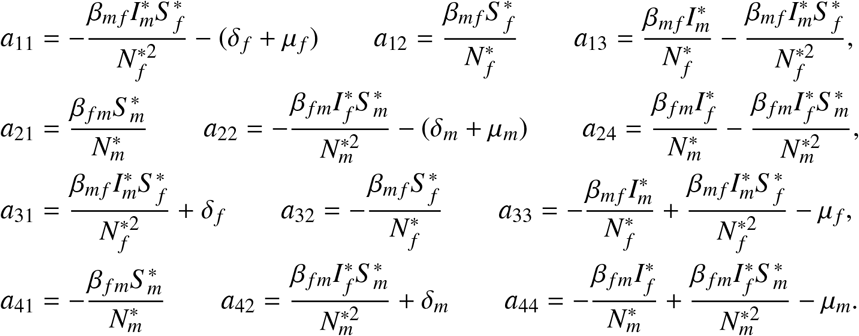

Then we have

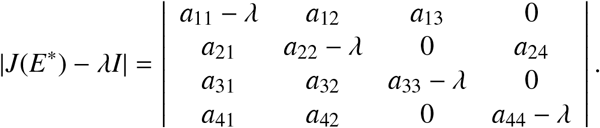

Adding the first row to the third row, and the second row to the last row, we get

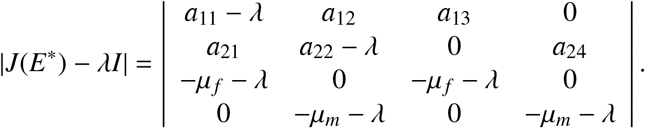

The characteristic equation is

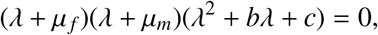

where

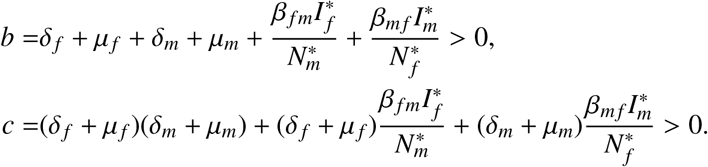

Clearly, all eigenvalues of *J*(*E*^*^) have negative real parts. Therefore, the endemic equilibrium *E*^*^ is locally asymptotically stable when *R*_0_ > 1.

## Appendix D Proof of Theorem 5

Considering limiting system, namely, 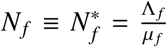 and 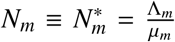, system (2) can be reduced to

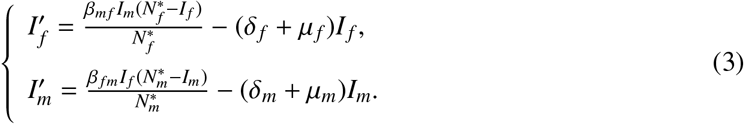

This is a two-dimensional system, to which we can apply Dulac’s criterion. We define

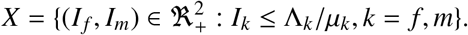

Denote

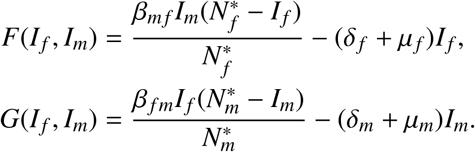

Using 1 as the Dulac multiplier, we get

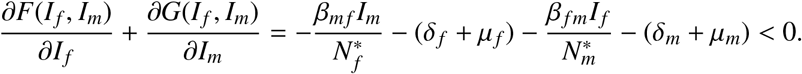

Therefore, there are no periodic orbits in region *X* [61]. Since the unique endemic equilibrium *E*^*^ is locally asymptotically stable when *R*_0_ > 1, it is globally asymptotically stable when *R*_0_ > 1.

## Appendix E Proof of Theorem 7

To get the endemic equilibrium, we set the right-hand side of system (1) to zero. At the equilibrium, we use 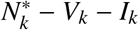 to replace *S* _*k*_, where 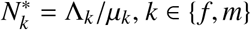. It follows that

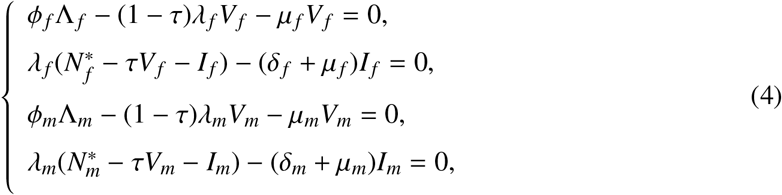

where

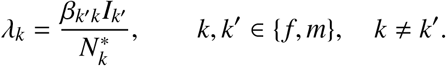

From the third equation of (4), we get

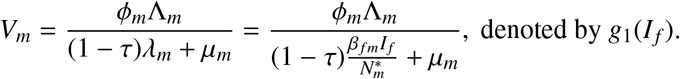

From the last equation of (4), we have *I*_*m*_ = *g*_2_(*I*_*f*_)*I*_*f*_, where

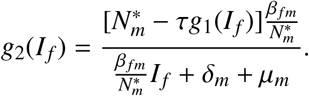

Substituting into the first equation of (4), we get

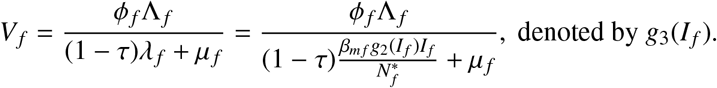

Substituting into the second equation of (4), we have

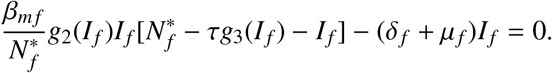

Define

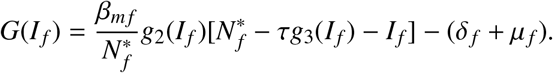

We notice that

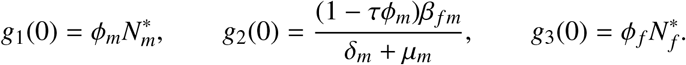

Hence

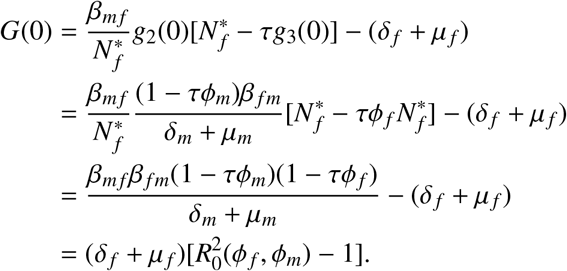

Thus, *G*(0) > 0 when *R*_0_(*ϕ*_*f*_, *ϕ*_*m*_) > 1. On the other hand,

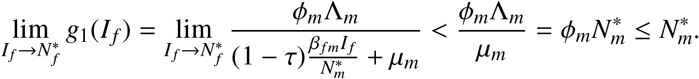

Hence, 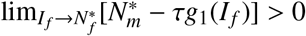. Consequently

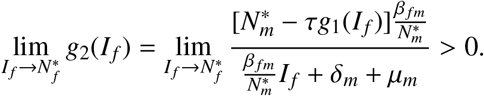

It follows that

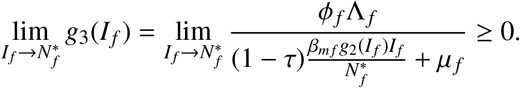

Therefore,

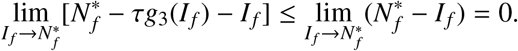

Since 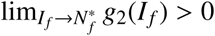,

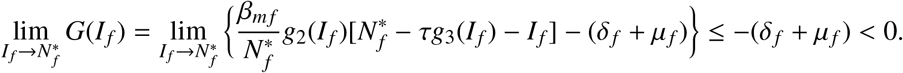

Therefore, there exists 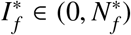 such that 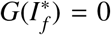. It follows that 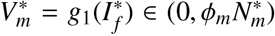, which indicates 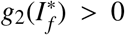. Hence 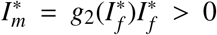. Since 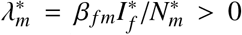, we have 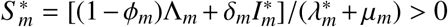. Since 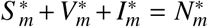, we have 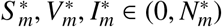. Similarly, we have 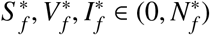. This completes the proof.

## Appendix F Proof of Theorem 8

From *ϕ*_*f*_ Λ_*f*_ + *ϕ*_*m*_Λ_*m*_ = *v*, we derive 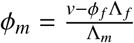. It follows that

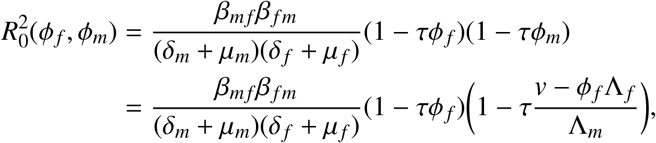

where

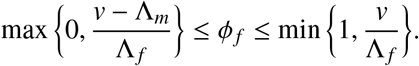

We have 4 cases.

**Case 1**. *v* ≤ Λ_*f*_ and *v* ≤ Λ_*m*_.

It is easy to check that

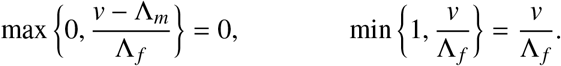

Hence

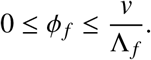

The value 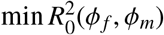 can only be attained at point *ϕ*_*f*_ = 0 or 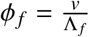. Since

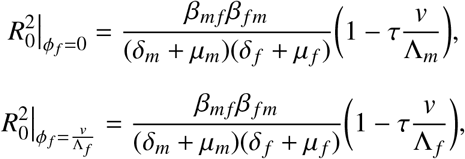

and min *R*_0_ and 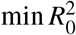 are obtained at the same point, we have the following results:

a. When *v* ≤ Λ_*m*_ < Λ_*f*_, min *R*_*0*_ (*ϕ*_*f*_, *ϕ*_*m*_) is attained at point 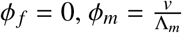;
b. When *v* ≤ Λ_*f*_ < Λ_*m*_, min *R*_0_(*ϕ*_*f*_, *ϕ*_*m*_) is attained at point 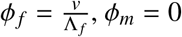;
c. When *v* ≤ Λ_*f*_ = Λ_*m*,_ min *R*_0_(*ϕ*_*f*_, *ϕ*_*m*_) is attained at point *ϕ*_*f*_ = 0, 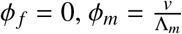 or 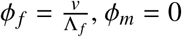;

**Case 2**. Λ_*f*_ < *v* ≤ Λ_*m*_.

In this case, we have

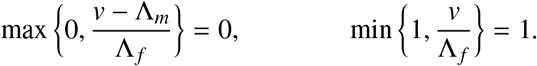

Thus, 0 ≤ *ϕ*_*f*_ ≤ 1. By the same reason as above, we compare

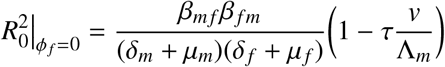

and

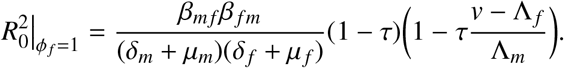

Since Λ_*f*_ < *v* ≤ Λ_*m*_ and *v* ≤ Λ_*f*_ + Λ_*m*_, we have 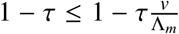 and 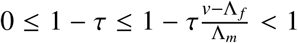, which implies 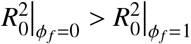. Therefore, we have the following result:

(d) When Λ_*f*_ < *v* ≤ Λ_*m*_, min *R*_0_(*ϕ*_*f*_, *ϕ*_*m*_) is attained at point 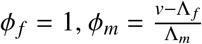.

**Case 3**. Λ_*m*_ < *v* ≤ Λ_*f*_.

By similar argument as in Case 2, we can derive the following result:

(e) When Λ_*m*_ < *v* ≤ Λ_*f*_, min *R*_0_(*ϕ*_*f*_, *ϕ*_*m*_) is attained at point 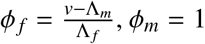.

**Case 4**. *v* > Λ_*f*_ and *v* > Λ_*m*_.

By similar argument as above, we compare

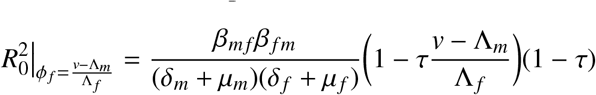

and

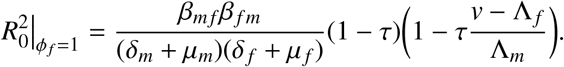

When Λ_*f*_ < Λ_*m*_, from *v* ≤ Λ_*f*_ + Λ_*m*_ we have 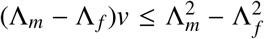. Hence 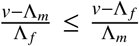. This implies 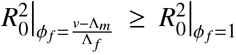. Similarly, when Λ_*f*_ > Λ_*m*_, we can derive 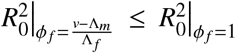.

Therefore, we have the following results:

(f) When Λ_*f*_ < Λ_*m*_ < *v*, min *R*_0_(*ϕ*_*f*_, *ϕ*_*m*_) is attained at point 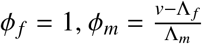 ;
(g) When Λ_*m*_ < Λ_*f*_ < *v*, min *R*_0_(*ϕ*_*f*_, *ϕ*_*m*_) is attained at point 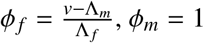;
(h) When Λ_*f*_ = Λ_*m*_ < *v*, min *R*_0_(*ϕ*_*f*_, *ϕ*_*m*_) is attained at point 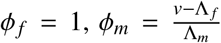 or 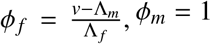.

Summing up all the results from (a)-(h), we get the following results. If Λ_*k*_ ≤ Λ_*k*_, min *R*_0_(*ϕ*_*f*_, *ϕ*_*m*_) is attained at: (i) 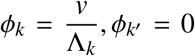 when *v* ≤ Λ_*k*_; (ii) 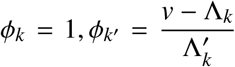 when *v* > Λ_*k*_, where *k, k ′* ∈ { *f, m*} and *k ≠ k ′*.

Considering the special case Λ_*f*_ = Λ_*m*_ = Λ, we only have Case 1 and Case 4. If we consider 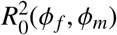 as a function of *ϕ*_*f*_, then it becomes a quadratic function, which has the axis of symmetry 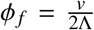. In either case, from *v* ≤ 2Λ we have 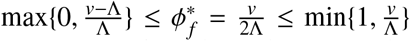. According to the properties of quadratic functions, max *R*_0_(*ϕ*_*f*_, *ϕ*_*m*_) can be achieved at 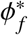, and the closer *ϕ*_*f*_ approaches 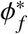, the bigger *R*_0_ is. Moreover, 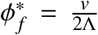 corresponds to 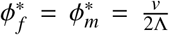. When *ϕ*_*f*_ approaches 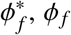 and *ϕ*_*m*_ are getting closer to each other. In other words, when Λ_*f*_ = Λ_*m*_ = Λ, max *R*_0_(*ϕ*_*f*_, *ϕ*_*m*_) is attained at 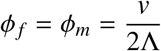, and the smaller |*ϕ*_*f*_ − *ϕ*_*m*_|, the bigger *R*_0_(*ϕ*_*f*_, *ϕ*_*m*_).

## Appendix G Proof of Corollary 1

Considering 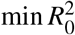 as a function of the vaccine amount *v* and denote it by *h*(*v*), we can summarize the results from Appendix F as follows:

i. When Λ_*f*_ ≤ Λ_*m*_

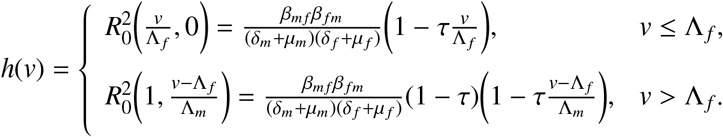
ii. When Λ_*f*_ > Λ_*m*_

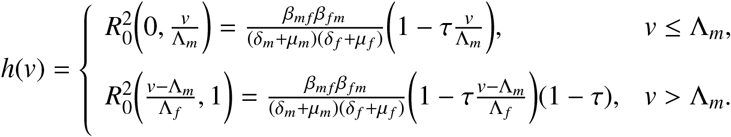

For Case (i), the derivative is

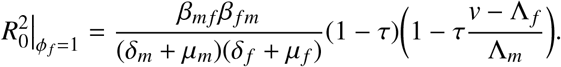

We have *h ′* (*v*) < 0. Since Λ_*f*_ ≤ Λ_*m*_, we have 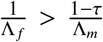. Therefore, | *h ′* (*v*) | is smaller for *v* ≤ Λ_*f*_ than that for *v* > Λ_*f*_. This shows that *h*(*v*) is a decreasing function and the decline speed when *v* ≤ Λ_*f*_ is greater than that when *v* > Λ_*f*_. This means that the vaccination becomes less effective once vaccine shots exceed the number of female recruits. A similar conclusion can be drawn for Case (ii).

## Appendix H Estimation of the numbers of 14-year-old boys and girls in Guangxi in 2021-2033

Under-five mortality rate (U5MR) is the probability of dying by age 5 per 1000 live births. The probability of dying among children aged 5-14 is about 18% of U5MR in the same year [62, 63]. Therefore, we estimate the number of children who are alive by age 14 to be newborn*(1-U5MR-U5MR*18%). The national census is only conducted at the year ending with 0, and the 1% national sample census is conducted at the year ending with 5. According to [64, 65], the sex ratio (male/female) is 115.6 (assume females are 100) for newborns in 1995 in China, and it is 112.06 for 15-year-old children in 2010. Thus, we estimate the sex ratio at age 14 to be the sex ratio at newborn (same cohort)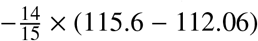. Using the number of 14 year-old children and the sex ratio for age 14, we can estimate the numbers of 14-year-old girls and boys in 2021-2033. The results are given in Table 8.

**Table 8:** Estimation of the numbers of 14-year-old girls and boys in 2021-2033 from newborns in 2007-2019*.

| Year | Newborn | U5MR | Sex ratio<br>(newborn) | Year | 14-year<br>children | Sex ratio<br>(age 14) | 14-year<br>girls | 14-year<br>boys |
| --- | --- | --- | --- | --- | --- | --- | --- | --- |
| 2007 | 710000 | 15.77 | 120.68 | 2021 | 696787.9 | 117.376 | 326623 | 383377 |
| 2008 | 720000 | 13.4 | 121.12 | 2022 | 708615.4 | 117.816 | 330554 | 389446 |
| 2009 | 720000 | 11.22 | 121.56 | 2023 | 710467.5 | 118.256 | 329888 | 390112 |
| 2010 | 720000 | 10.88 | 122 | 2024 | 710756.4 | 118.696 | 329224 | 390776 |
| 2011 | 710000 | 9.7 | 120.4 | 2025 | 701873.3 | 117.096 | 327044 | 382956 |
| 2012 | 740000 | 7.89 | 118.8 | 2026 | 733110.5 | 115.496 | 343394 | 396606 |
| 2013 | 750000 | 7.73 | 117.2 | 2027 | 743159 | 113.896 | 350638 | 399362 |
| 2014 | 720000 | 7.74 | 115.6 | 2028 | 713424.1 | 112.296 | 339149 | 380851 |
| 2015 | 720000 | 6.25 | 114 | 2029 | 714690 | 110.696 | 341725 | 378275 |
| 2016 | 770000 | 6.03 | 113.446 | 2030 | 764521.1 | 110.142 | 366419 | 403581 |
| 2017 | 820000 | 4.99 | 112.892 | 2031 | 815171.7 | 109.588 | 391244 | 428756 |
| 2018 | 710000 | 4.98 | 112.338 | 2032 | 705827.8 | 109.034 | 339658 | 370342 |
| 2019 | 660000 | 4.77 | 111.23 | 2033 | 656285.1 | 107.926 | 317421 | 342579 |

\*U5MR: under 5 mortality rate; Sex ratio: male/female (female is 100); The number of 14-year children is derived from the number of newborn and U5MR; Sex ratio at age 14 is derived from sex ratio at newborn; Numbers of 14-years girls and boys are derived from the number of 14-year children and the sex ratio at age 14 (for details see Appendix H).

## Acknowledgments

This work was supported by the NSF grants DMS-1951595 (MM), DMS-1620957 (MH) and DMS-1950254 (LR).

## Notes

### Competing Interest Statement

The authors have declared no competing interest.

### Funding Statement

This study was funded the NSF grants DMS-1951595 (MM), DMS-1620957 (MH) and DMS-1950254 (LR).

